# Trends of Cutaneous Leishmaniasis, Western Ethiopia: retrospective study

**DOI:** 10.1101/2022.04.09.22273646

**Authors:** Zalalem Kaba Babure, Yusuf Mohammed Ahmed, Getu Mosisa

## Abstract

**Background:** Cutaneous leishmaniasis (CL) is the most common form of leishmaniasis and causes skin lesions, mainly ulcers, on exposed parts of the body, leaving life-long scars and serious disability or stigma. In Ethiopia, cutaneous leishmaniasis is primarily caused by Leishmania aethiopica and less often by Leishmania Tropica and Leishmania major. There is a major prevalence gap in study areas. Hence, this study assessed the trends of cutaneous leishmaniasis in the western part of Ethiopia.

**Methodology:** A three-year retrospective study (09 October 2018 to 31 January 2022) was conducted by extracting information from the national leishmaniasis register for patients visiting the Nekemte Specialized Hospital (NSH) treatment center, Nekemte, Western Ethiopia. A standard data abstraction checklist was used to review Leishmaniasis records. Data were extracted from national leishmaniasis cases registration book by principal investigators and summarized using Microsoft Excel. All data were entered and analyzed using the Excel Microsoft office package.

**Results:** A total of 64 patients were treated for cutaneous leishmaniasis in the area during the study period. About 35(54.69%) cutaneous leishmaniasis cases were males, and the median age for sex was 18.5 years. Most of the cases were among those aged 15-24 years (39.1%) while extreme age groups reported the least. About 35 (54.69%) of cutaneous leishmaniasis cases were from rural areas, and two-thirds (31, 65.96%) of patients were seeking of medical treatment after 3-6 months developing sign and symptoms. One-fourth (17, 26.56%) of CL cases were reported in January followed by August (10, 15.63), and there were no cases reported in June and October.

**Conclusion:** The most affected age group are those 15-24 years and those from rural communities. January is months most cases reported and late coming to treatment and needs awareness creation.

**Author summary:** Globally, cutaneous leishmaniasis (CL) is the most common form of leishmaniasis which accounts for about 95% of cases. It is an emerging uncontrolled and neglected infection affecting millions yearly. Most CL patients are residing in low- to middle-income countries, where limited healthcare budgets and a large burden caused by other ailments such as malaria, tuberculosis, and HIV (human immunodeficiency virus) are prominent. Accurate disease burden is challenging since misdiagnosis is common, and there are no standard reporting guidelines. There is limited information regarding the magnitude of the cases in low and middle-income countries, including Ethiopia. The lack of epidemiological burden and distribution makes it difficult to advocate for control activities and further research to inform public health policy. This study aimed to assess the trends of CL in the western part of Ethiopia, to fill the gaps in the dearth of information in the area. The study highlighted the distribution of CL cases by gender, age, seasons of the year, and geographical areas (rural or rural). Moreover, we recommend community-based research programs to determine the exact incidence and prevalence of CL cases and associated risk factors in the western part of Ethiopia, particularly in the East Wollega Zone.

## Introduction

Leishmaniasis is a parasitic protozoan transmitted to humans by the bite of infected female sand flies. It causes three main forms of leishmaniasis: visceral (the most serious form of the disease), cutaneous (the most common form), and mucocutaneous. Cutaneous leishmaniasis (CL), the most common form, accounts for about 95% of cases globally [1]. CL is an emerging uncontrolled and neglected infection affecting millions yearly. Most CL patients are living in low- to middle-income countries where limited healthcare budgets and a large burden caused by other ailments such as malaria, tuberculosis, and HIV (human immunodeficiency virus are prominent [2].

CL is the most common form of leishmaniasis and causes skin lesions, mainly ulcers, on exposed parts of the body, leaving life-long scars and serious disability or stigma. It is estimated that between 600,000 to 1 million new cases occur worldwide annually [3]. It is endemic in the tropics and neotropics. Despite its increasing worldwide incidence, but because it is rarely fatal, CL has become one of the so-called neglected diseases, with little interest by financial donors, public-health authorities, and professionals to implement activities to research, prevent, or control the disease [4]. It presents in 67 countries in the old World (Europe, Africa, Middle East, Central Asia, and the Indian subcontinent). When susceptible populations become exposed, CL may result in noticeable epidemics, such as in Burkina Faso, Ghana and new pockets in Ethiopia. CL is rarely fatal but it can cause substantial suffering because of the related stigma and the disfiguring scars it leaves in a number of cases. Accurate disease burden is challenging since misdiagnosis is common and there are no standard reporting guidelines. There is limited information regarding the magnitude of the cases in low and middle-income countries, including Ethiopia. The lack of epidemiological burden and distribution makes it difficult to advocate for control activities and further research to inform public health policy [5].

The ten countries with the highest estimated CL case counts, Afghanistan, Algeria, Colombia, Brazil, Iran, Syria, Ethiopia, North Sudan, Costa Rica, and Peru, together account for 70 to 75% of global estimated CL incidence. Approximately, 0.7 to 1.2 million CL cases occur each year, globally [6]. In Ethiopia, CL is primarily caused by Leishmania aethiopica and less often by Leishmania Tropica and Leishmania major. Leishmania aethiopica causes both diffuse cutaneous leishmaniasis (DCL) and localized cutaneous leishmaniasis (LCL), which are found in the highlands of Ethiopia. Leishmania aethiopica occurs only in the Ethiopian and Kenyan highlands and its reservoir is the rock hyrax, while the vector is P. larroussius[7-9].

The study conducted in ALERT (All African Leprosy Rehabilitation and Training Center) Hospital, Addis Ababa, Ethiopia showed that based on geographical areas of the patients’ flow Addis Ababa (41%, 96/234) and Oromia (30.3%, 71/234) had highest leishmaniasis distribution. Besides, it revealed that among 71 CL cases reported from Oromia region, 7(9.9%) of them were from Wollega [10]. However, there is no study conducted in four Wollega Zones and western part of the country which reveals the trends of CLs. Hence, this assessment used to fill the gaps in dearth of information.

## Methods and Materials

### Study design and settings

A three-year retrospective study was conducted by extracting information from the national leishmaniasis register for patients visiting the Nekemte Specialized Hospital (NSH) treatment center between October 09, 2018, and January 31, 2022, Nekemte, Western Ethiopia. Based on the reviewed records, patients were coming to NSH treatment center from different Zones and districts of Western Ethiopia. The study was conducted at NSH CL treatment center, located in Nekemte town, Western Ethiopia. This center provides services for patients coming from 17 districts of East Wollega Zone and neighbouring Zones like Horo Guduru, West Wollega, West Shoa, Buno Bedele, Ilu-ababor, and Kellem Wollega in western part of the country.

### Study population and selection criteria

We conducted a retrospective analysis of CL patients’ medical records at NSH treatment center between October 09, 2018, and January 31, 2022. The extracted data include demographic data; diagnostic laboratory results and clinical information that were collected and assessed from a total of 64 cutaneous leishmaniasis cases. Each data element was carefully collected to avoid redundancy.

### Data collection method and tools

Demographic (sex, age, and residence) and clinical (duration of illness/sick before admissions, size of lesions, and treatment outcome) information, and laboratory results (parasitological and HIV testing) on all registered CL cases are systematically collected using standardized data abstraction forms. A standard data abstraction checklist was used to review NSH treatment center Leishmaniasis records from October 09, 2018, to January 31, 2022. Leishmaniasis register hospital records were reviewed by principal investigators.

### Data analysis and interpretation

This retrospective analysis included the analysis of the reported CL cases using the available hospital records at the NSH CL treatment center, in western Ethiopia. Data were extracted from the national leishmaniasis cases registration book and summarized using Microsoft Excel. All data were entered and analysed using the Excel Microsoft office package. Categorical data were expressed using frequencies, percentages, and proportions with trend analysis when applicable. For continuous data, the median was used for expression.

### Operational definition

#### Cutaneous Leishmaniasis

A protozoan infection and vector borne disease of human beings caused by Leishmania specious that affects the skin primarily [11].

#### Trends of Cutaneous Leishmaniasis

The occurrence and distribution of the disease in the population at a given period [11].

### Ethical consideration

Ethical approval was obtained from East Wollega Zonal Health Department, Public and Health Emergency Management (PHEM) case team institutional review board. Consent was also sought from the hospital administration before being involved. Since the data were extracted form secondary data; formal consent (verbal or written) from parent/guardian was not required. Besides, we used patient identifiable codes to maintain the confidentiality of each patient.

## Results

### Sociodemographic characteristics

Starting from October 09, 2018, to January 31, 2022, a total of 64 patients were treated for cutaneous Leishmaniasis (CL) at Nekemte Specialized Hospital treatment Centre. Of the total cases with CL, about 35(54.69%) were males and 29(45.31%) of them were females. The median age for sex was 18.5 years, while that of males and females were 19 years and 18 years, respectively. The maximum and minimum age for males was 3 years and 45 years, while that of females was 4 years and 50 years. The majority of the cases were from the Nekemte council (29, 45.31%), and East Wollega Zone (29, 45.31%). Among 17 districts found in East Wollega Zone, CL cases were reported from 9 districts. More than half of CL cases (17, 58.62%) were reported from three districts of the Zone: -Gida Ayana (4, 6.25%), Leka Dulecha (5, 7.81%), and Jima Arjo (8, 12.5%). CL affected all age categories from 3 years to 50 years, with a median age of 18.5 years. Most of the cases were those aged 15-24 years (39.1%), followed by 5-14 years (26.6%), while extreme age groups reported the least. More than half of CL cases (35, 54.69%) were from rural areas. About two-thirds (31, 65.96%) of patients were sick of medical treatment between 3-6 months, and in more than three-fourths (11, 78.57%) the size of the lesion was less than four centimetres [Table 1].

**Table 1:**
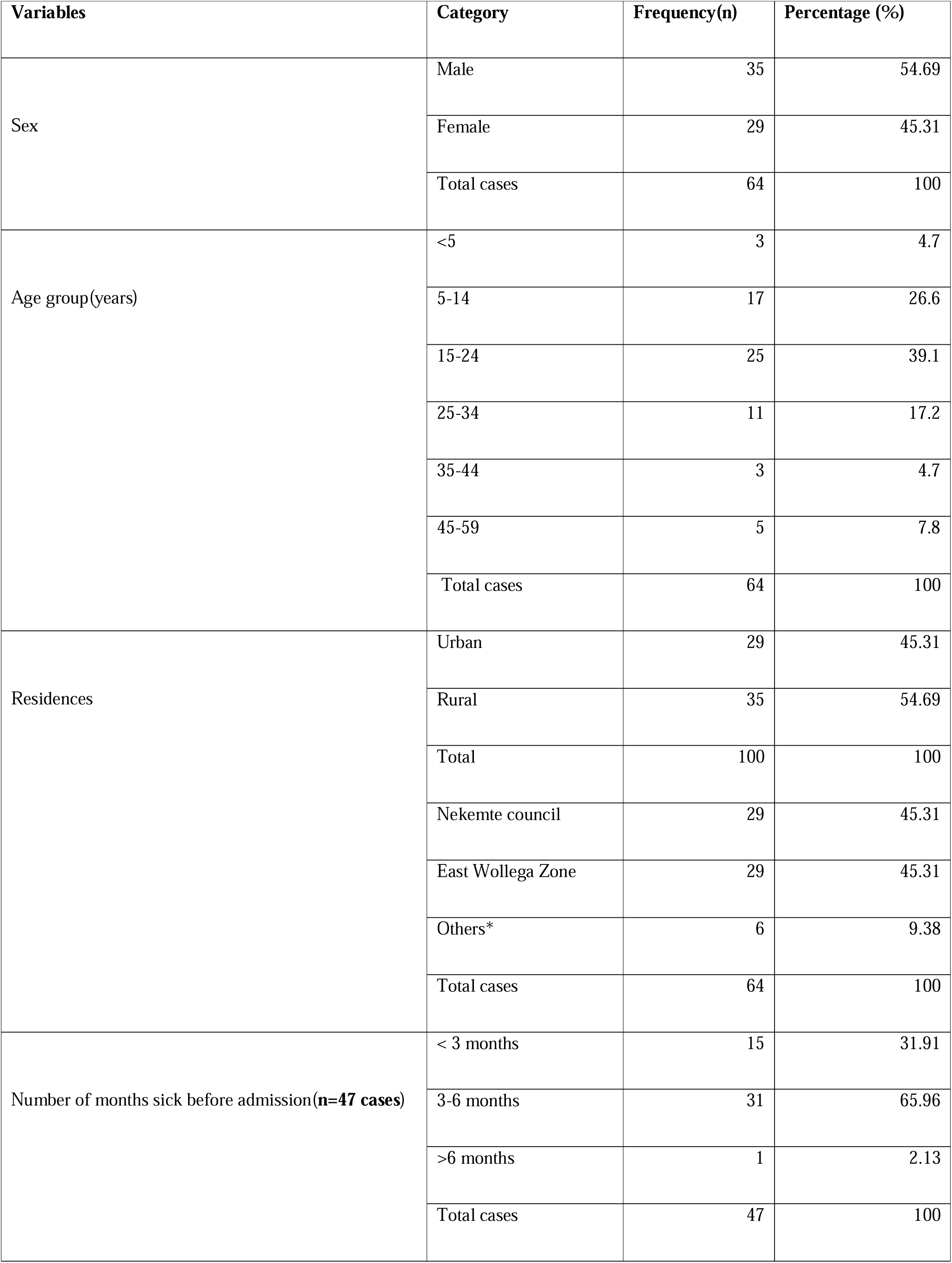
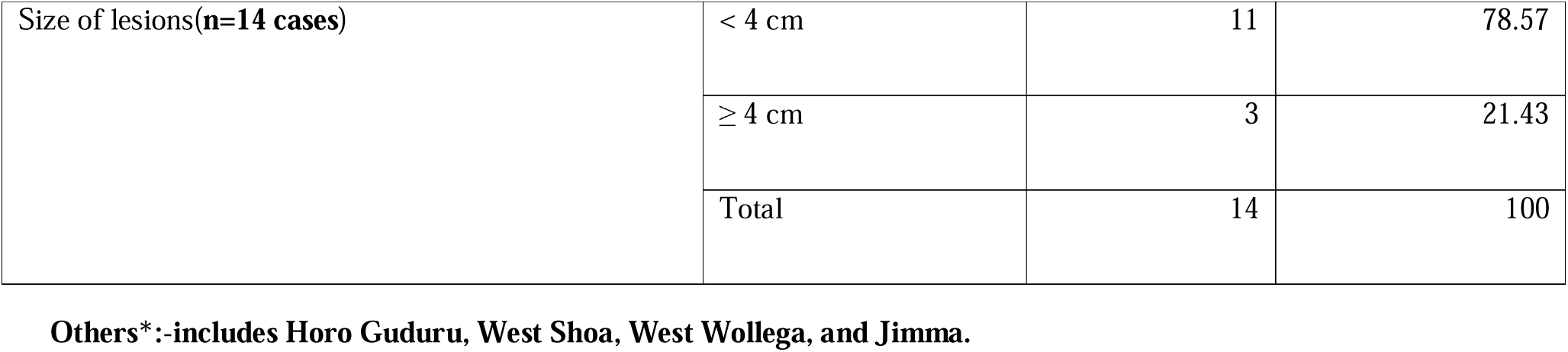
Demographic and clinical characteristics of patients with cutaneous leishmaniasis (CL) based on Nekemte Specialized Hospital CL treatment center record review, Western Ethiopia, 2022(n=64).

| Variables | Category | Frequency(n) | Percentage (%) |
| --- | --- | --- | --- |
| Sex | Male | 35 | 54.69 |
|  | Female | 29 | 45.31 |
|  | Total cases | 64 | 100 |
| Age group(years) | <5 | 3 | 4.7 |
|  | 5-14 | 17 | 26.6 |
|  | 15-24 | 25 | 39.1 |
|  | 25-34 | 11 | 17.2 |
|  | 35-44 | 3 | 4.7 |
|  | 45-59 | 5 | 7.8 |
|  | Total cases | 64 | 100 |
| Residences | Urban | 29 | 45.31 |
|  | Rural | 35 | 54.69 |
|  | Total | 100 | 100 |
|  | Nekemte council | 29 | 45.31 |
|  | East Wollega Zone | 29 | 45.31 |
|  | Others* | 6 | 9.38 |
|  | Total cases | 64 | 100 |
| Number of months sick before admission(**n=47 cases**) | < 3 months | 15 | 31.91 |
|  | 3-6 months | 31 | 65.96 |
|  | >6 months | 1 | 2.13 |
|  | Total cases | 47 | 100 |
| Size of lesions(n=14 cases) | < 4 cm | 11 | 78.57 |
|  | ≥ 4 cm | 3 | 21.43 |
|  | Total | 14 | 100 |

**Others\*:-includes Horo Guduru, West Shoa, West Wollega, and Jimma.**

### Age and Sex distribution of CL cases

This assessment indicated that young children and adolescents (15-34 years) of school-going age were disproportionately affected (37, 57.81%). The sex structure of patients with cutaneous leishmaniasis shows an almost one-to-one ratio (1.21: 0.81) [Table 2].

**Table 2:** Age and sex distribution of patients with Cutaneous Leishmaniasis at Nekemte Specialized Hospital treatment center from October 09, 2018 to January 31, 2022, Nekemte, Western Ethiopia, 2022(n=64).

| Age group(years) | Sex category |  |  |  |  |  |
| --- | --- | --- | --- | --- | --- | --- |
|  | Male | % | Female | % | Total | Percentage (%) |
| <5 | 2 | 66.67 | 1 | 33.33 | 3 | 4.69 |
| 5-14 | 8 | 50 | 8 | 50 | 16 | 25 |
| 15-24 | 13 | 50 | 13 | 50 | 26 | 40.63 |
| 25-34 | 9 | 81.82 | 2 | 18.18 | 11 | 17.19 |
| 35-44 | 1 | 33.33 | 2 | 66.67 | 3 | 4.69 |
| 45-59 | 2 | 40 | 3 | 60 | 5 | 7.81 |
| **Total** | **35** | **54.69** | **29** | **45.31** | **64** | **100** |

#### Distribution of CL cases among East Wollega Zone Districts

Among the total 64 CL cases reviewed from the hospital record, about (29, 45.31%) of CL cases were from East Wollega Zone districts. Among 17 districts found in East Wollega Zone, CL cases were reported from 9 of them. More than half of CL cases (17, 58.62%) were reported from three districts of the Zone: -Gida Ayana (4, 6.25%), Leka Dulecha (5, 17.24%), and Jima Arjo (8, 27.59%) [Figure 1].

**Figure 1:**
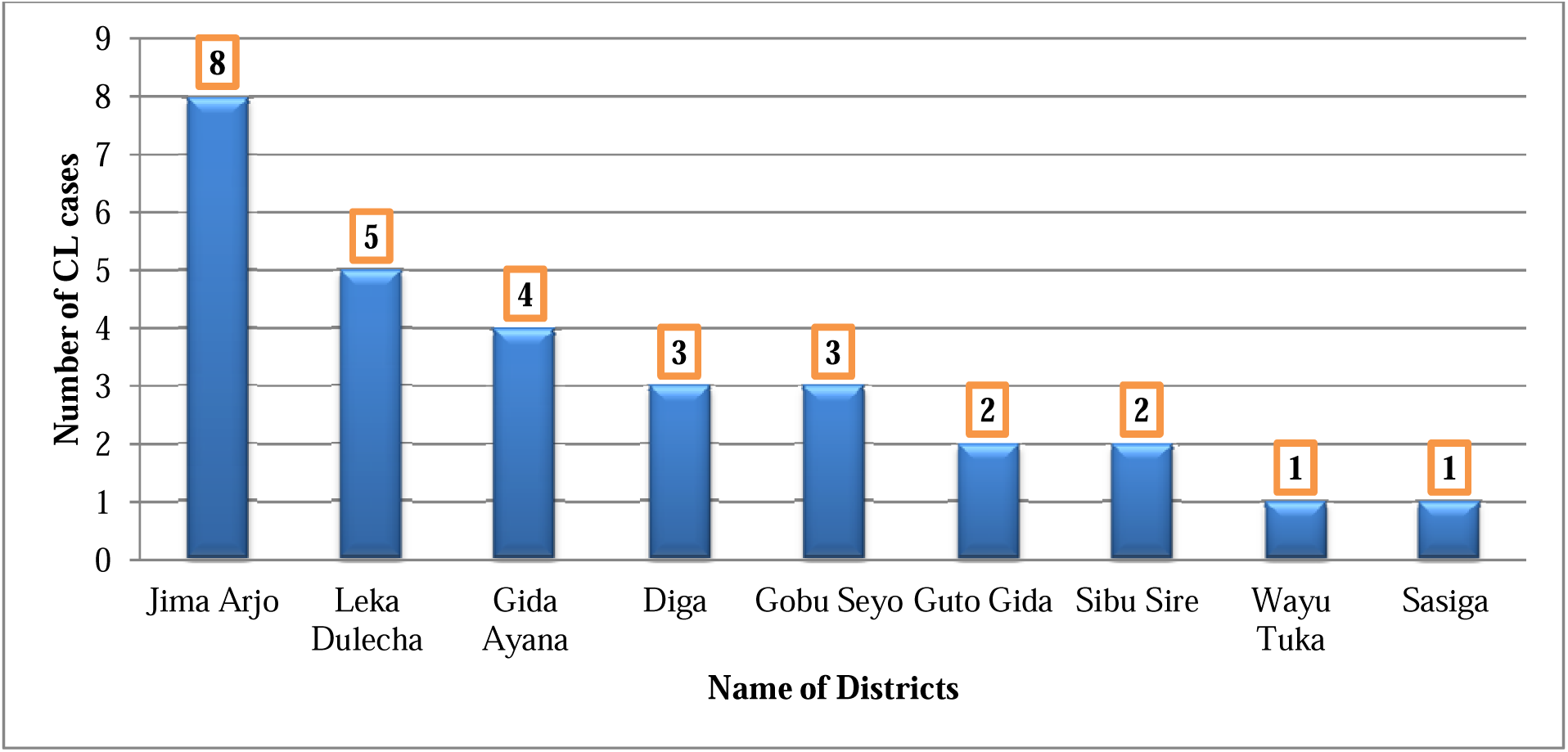
Distribution of CL cases by districts in East Wollega Zone, Nekemte, Western Ethiopia, 2022(n=29).

#### Distribution of cases by kebeles of Nekemte council

The study showed that nearly three-fourths (21, 72.41%) of cutaneous leishmaniasis cases were reported from Kebele 04 of Nekemte council. Besides, the least was reported from Kebele 01, 06, and 07[Figure 2].

**Figure 2:**
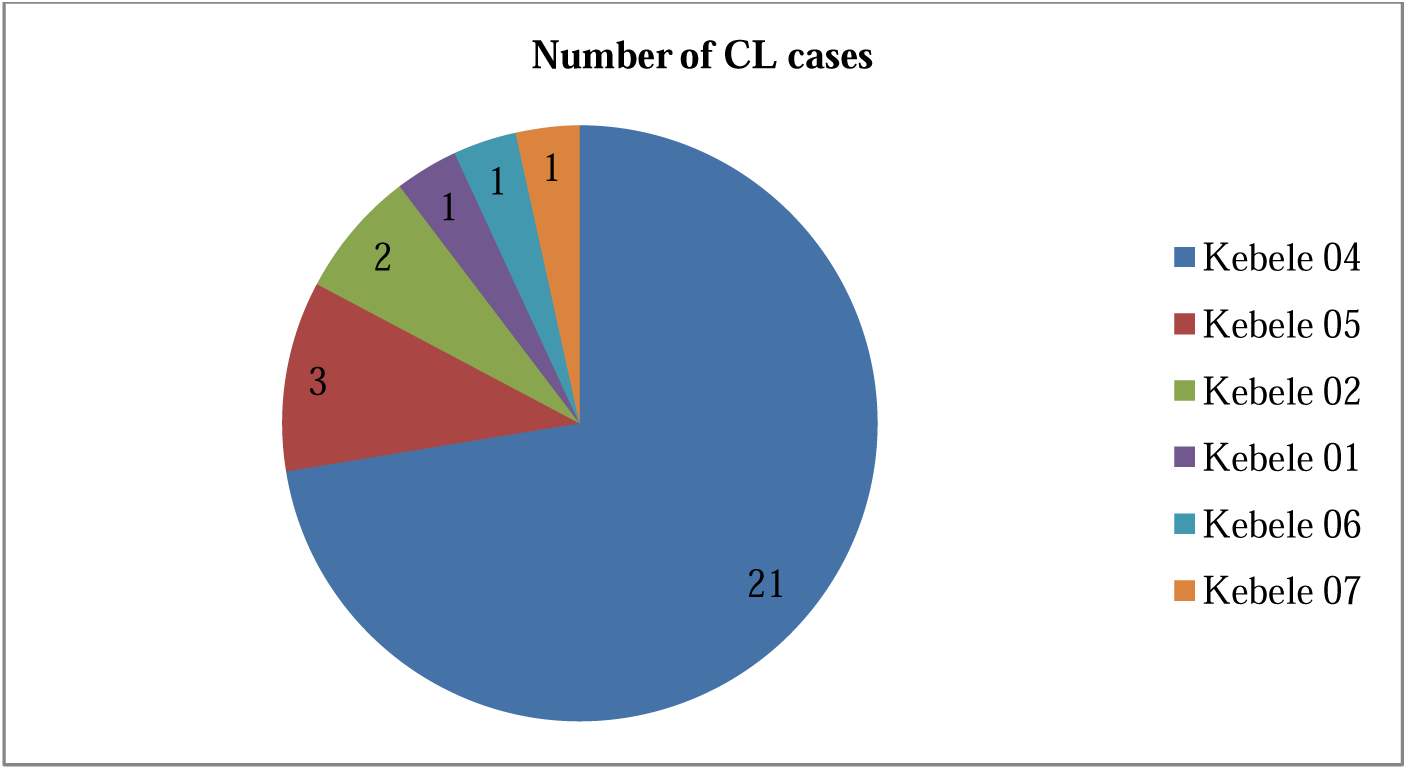
Distribution of CL cases by Kebeles in Nekemte council, Nekemte, Western Ethiopia, 2022(n=29).

#### Number of months’ sick before admission

The median number of months sick before admission was 3 months, while the minimum was 1 month and the maximum was 9 months. Nearly more than one-quarter of patients seeks care within 2 months (12, 25.53%) and 3 months (13, 27.66%) respectively. But, a significant number of patients (13, 27.66%) seek medical care greater than or equal to 5 months at Nekemte Specialized Hospital treatment center, Ethiopia. From those late medical treatment seekers, most of them (7/13, 53.85%) were from Nekemte town [Figure 3].

**Figure 3:**
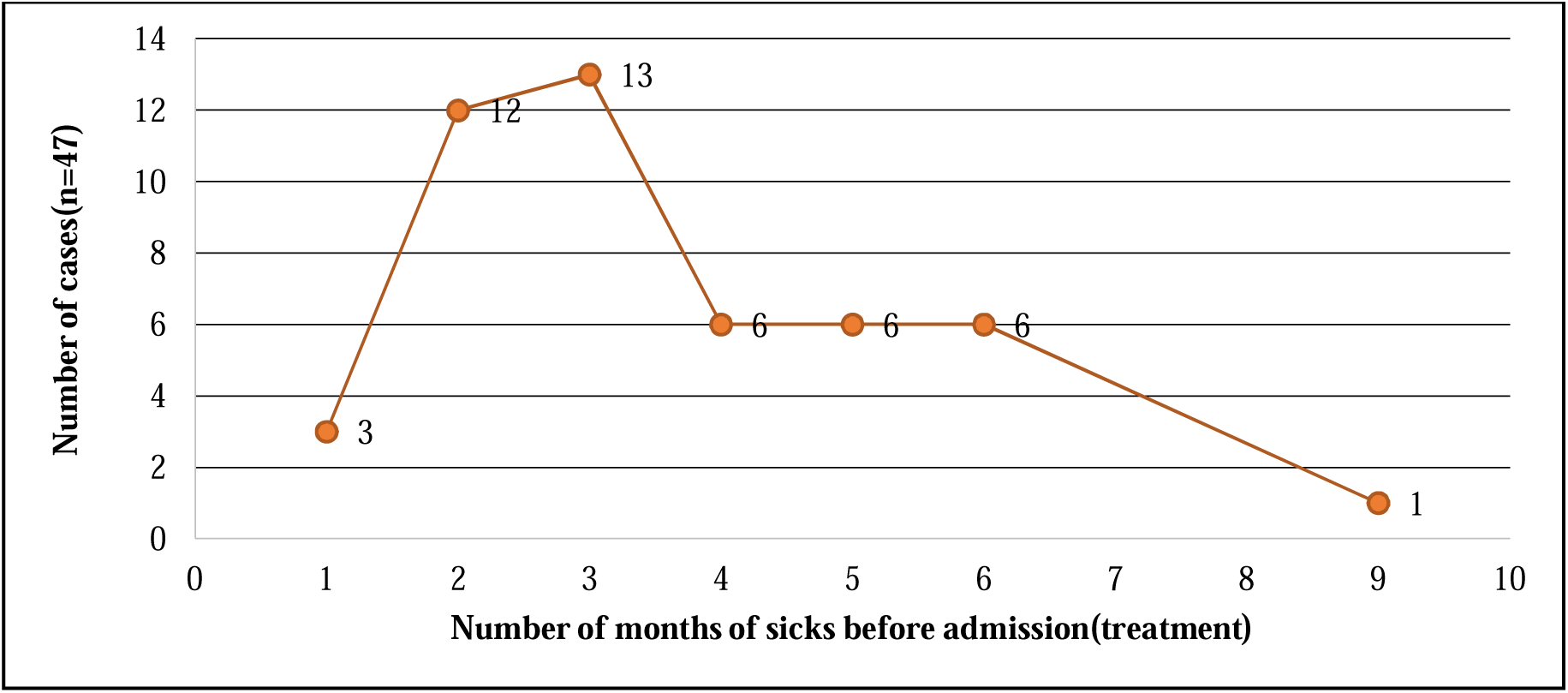
Line graph showing the distribution of cutaneous leishmaniasis cases by several months sick before admission based on record review, Nekemte, Western Ethiopia(n=47),2022.

#### Yearly trends of Cutaneous Leishmaniasis in Western Ethiopia

This assessment showed that the number of cutaneous leishmaniasis was slightly increased from 2018/2019 to 2020 and then declined from 2020 to 2021in the last more than 2 years. The total number of CL cases was 18, 20, 16, and 10 from October 09, 2018, to January 31, 2022, respectively [Figure 4].

**Figure 4:**
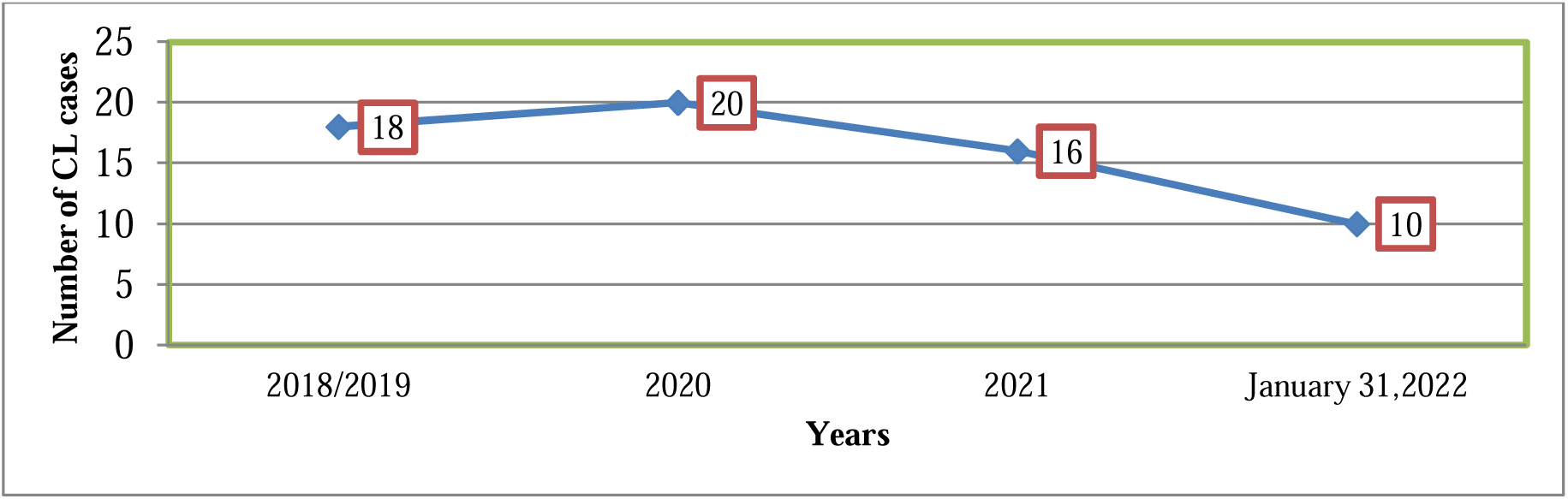
Line graph showing the trends of cutaneous leishmaniasis cases based on Nekemte Specialized Hospital treatment center record review, Nekemte, Western Ethiopia, 2022.

#### Quarterly trends of Cutaneous Leishmaniasis in Western Ethiopia

The study revealed that half (33, 51.56%) of CL cases were reported in quarter three (January, February, and March), followed by quarter one (July, August, and September) which was about (19, 29.69%) CL cases reported [Table 3].

**Table 3:**
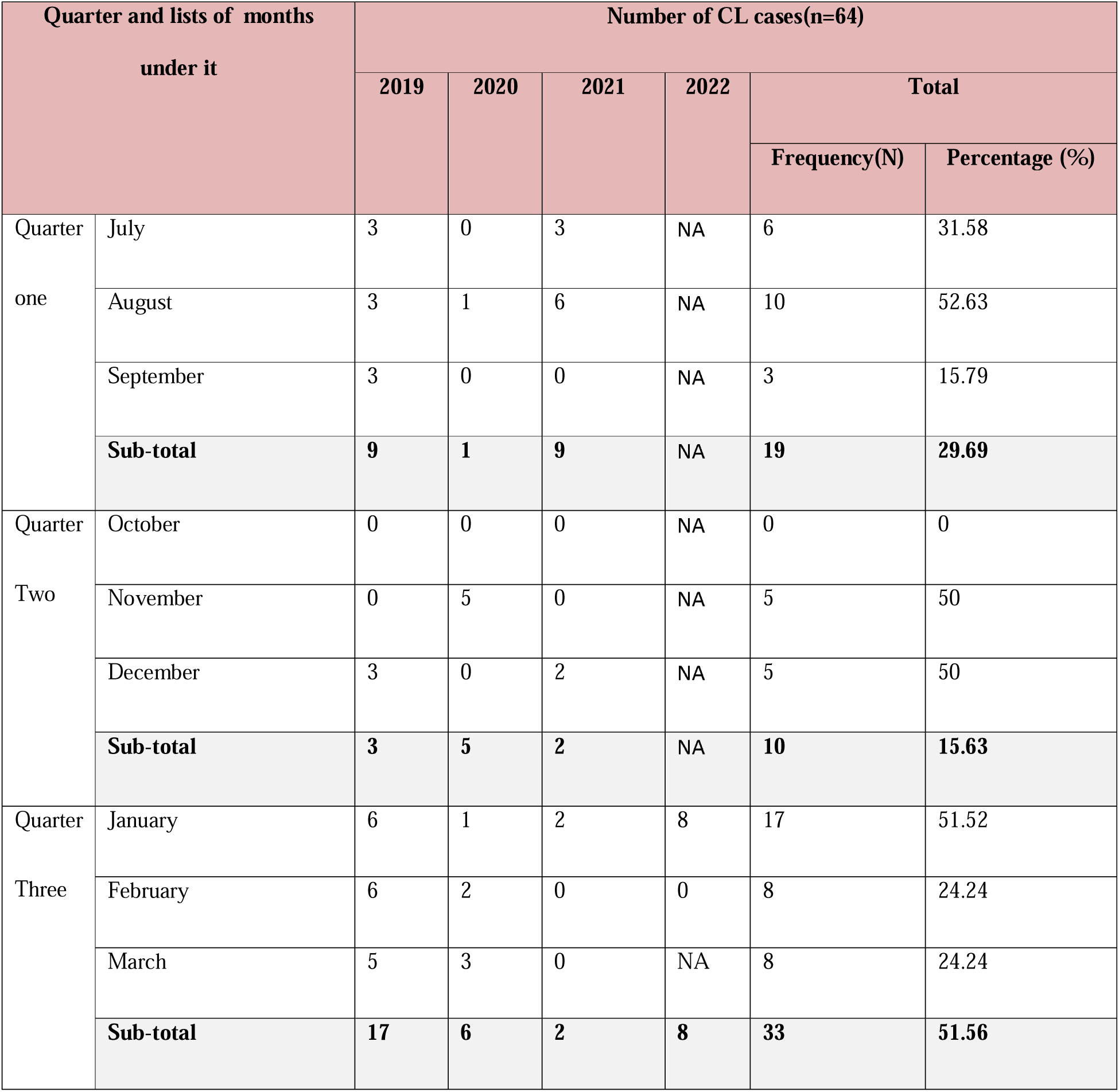
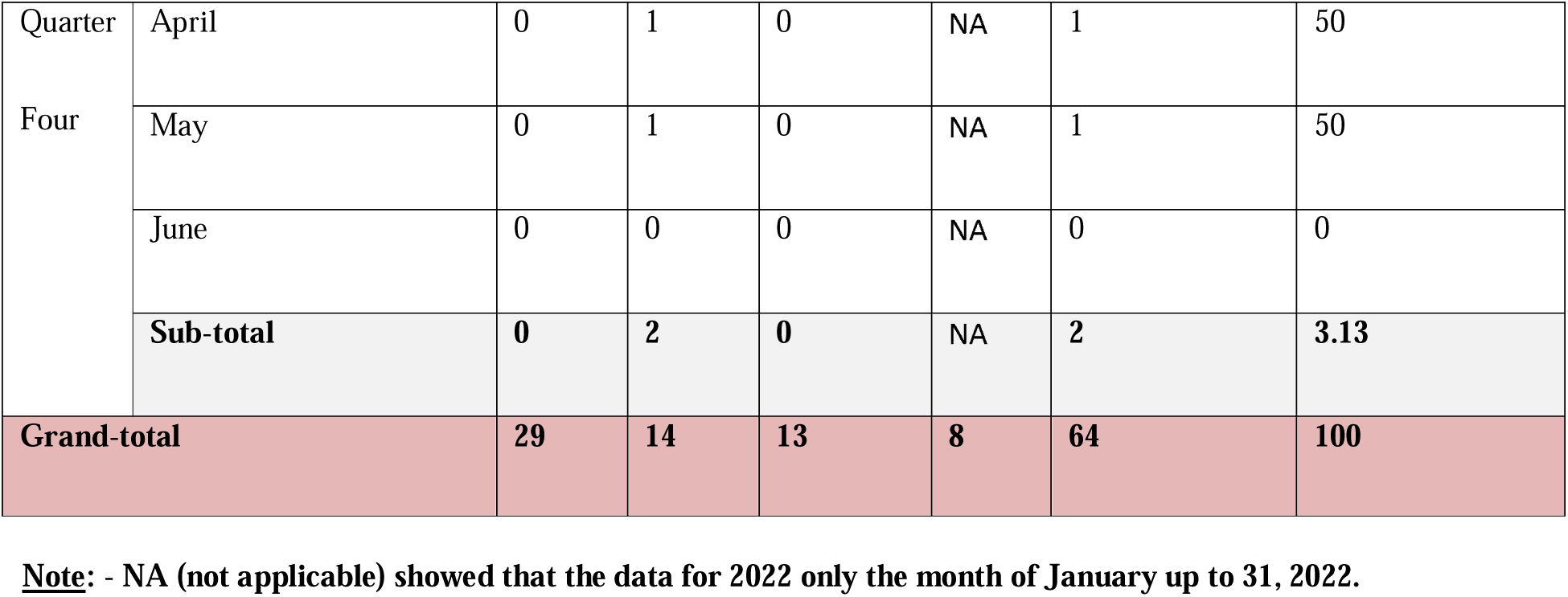
A Quarter trends of reported CL cases based on Nekemte Specialized Hospital treatment center record review, Western Ethiopia, 2022.

#### Cumulative monthly trends of Cutaneous Leishmaniasis in Western Ethiopia

The study revealed that one-fourth (17, 26.56%) of CL cases were reported in January followed by August (10, 15.63). Besides, there were no cases reported in June and October [Figure 5].

**Figure 5:**
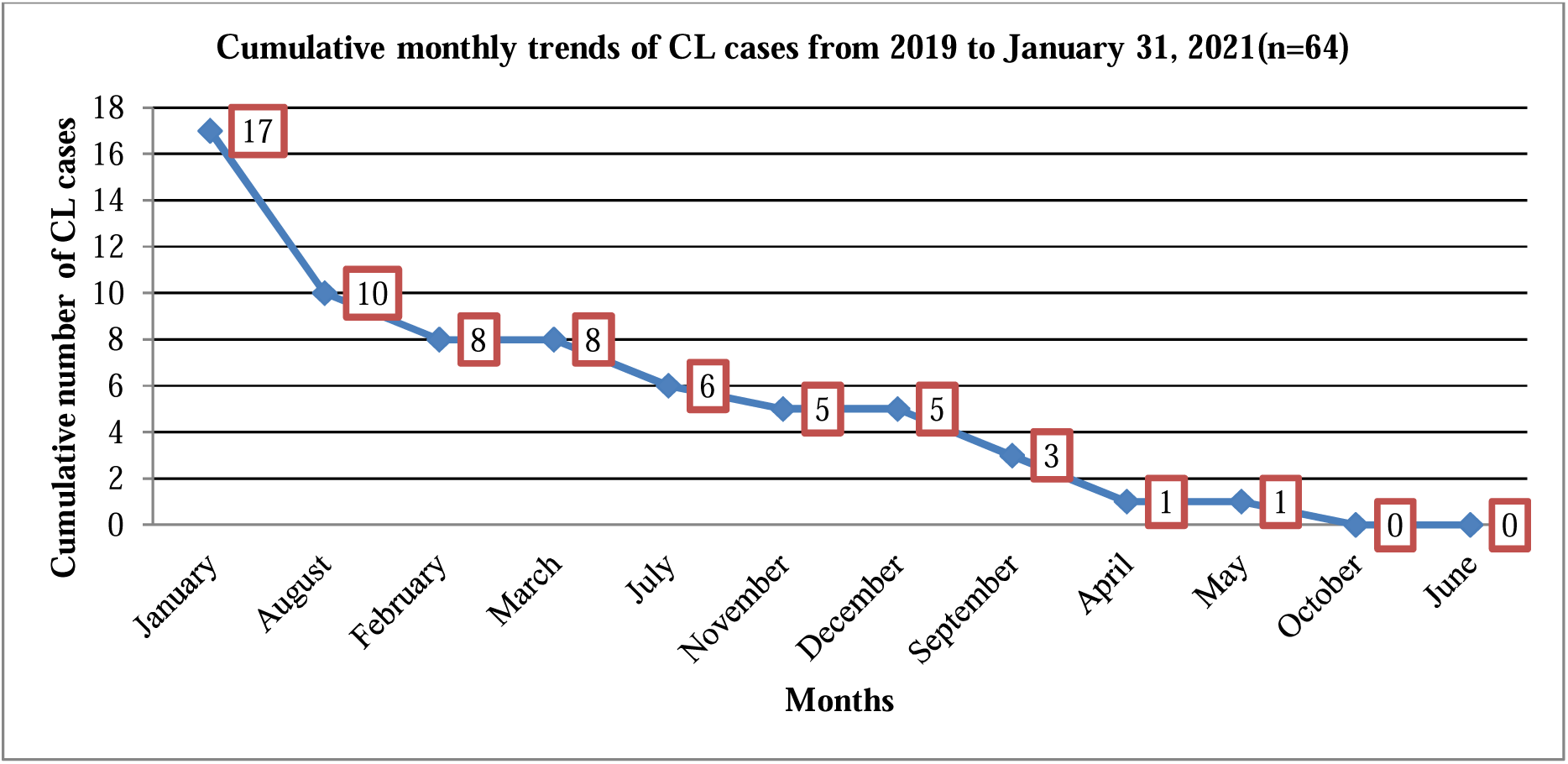
Line graph showing the cumulative monthly trends of cutaneous leishmaniasis cases based on Nekemte Specialized Hospital treatment center record review from 2019 to January 31, 2022, Nekemte, Western Ethiopia, 2022.

#### Monthly trends of Cutaneous Leishmaniasis in Western Ethiopia

The study revealed that there was a case build up in the months of July (6 cases), February (8 cases), March (8 cases), August (10 cases), and January (17 cases) [Figure 6].

**Figure 6:**
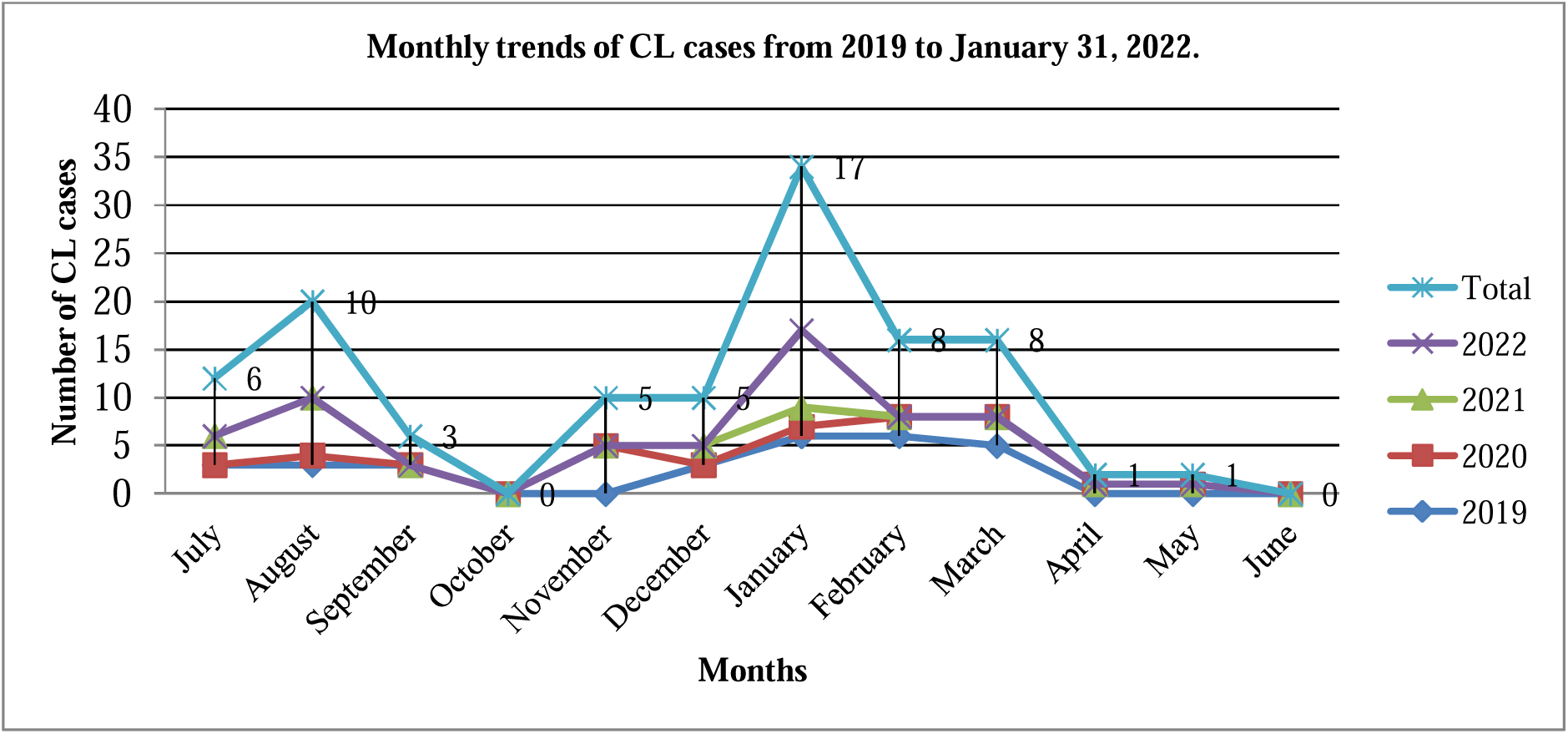
- Line graph showing the monthly trends of CL cases reported based on Nekemte Specialized Hospital treatment center record review from 2019 to January 31, 2022, Western Ethiopia. **<u>Note</u>**: - The date for the year 2022 was only up to January 31, 2022 and the rest of the months in the year filled zero (0) when sketching the line graph.

The study also indicated that all the patients were cured and there were no admitted cases. Besides, it showed that HIV testing was offered and performed for all the patients and there was no positive case found.

## Discussion

This study revealed that the trends of CL cases in western Ethiopia were fluctuating; it was slightly increased from 2018/2019 to 2020 and then declined from 2020 to 2021, which is in line with the ten-year trend analyses report in the University of Gondar Hospital, Northwest Ethiopia [12]. In contrast to this, the study conducted in Northeast Ethiopia revealed that the number of CL cases was increased significantly from year to year [11]. These could be due to the differences in socio demographic and economic characteristics of the populations.

Our study revealed that males were more affected by CL than females which agree with different studies [10-17]. However, our finding is analogized with the studies reported from Algeria, Istanbul, Turkey, and Larstan, South of Iran that excess in the frequency of CL cases among females than males [18-20]. In contrast to this, the studies conducted in rural communities in Tigray, northern Ethiopia, Silti woreda, Ethiopia, and Pakistan reported no gender difference in the frequency of CL cases [21-23]. The possible explanation for this is due to the high engagement of males in outdoor activities like farming and different agricultural works compared to females.

In this study, more than half of CL cases (35, 54.69%) were from rural areas which are incomparable with the study done in ALERT(All African Leprosy Rehabilitation and Training Center) Hospital, Addis Ababa, Ethiopia, and Larstan, South of Iran which showed that the distribution of CL patients was higher among rural than urban [10,20]. In contrast to this, the study conducted in Northeast Ethiopia, and in Silti woreda, Ethiopia revealed a higher distribution of CL cases in urban areas than in rural ones [11,22]. This variation could be due to the migration of infected CL cases from rural to urban and the higher prevalence of the cases in highland areas than in lowland areas of the study sites.

This study also revealed that most of the CL cases were among those aged 15-24 years (39.1%), followed by 5-14 years (26.6%). These indicated that young children and adolescents (15-34 years) of school-going age were disproportionately affected (37, 57.81%) by CL cases which is comparable with studies conducted in different areas [10, 16-17, 19, 21-23]. In this study, extreme age groups, less than 5years and greater than 45 years were the least affected by CL cases which is comparable with the study conducted at the University of Gondar Hospital, Northwest Ethiopia [12].

In this study, about two-thirds (31, 65.96%) of patients were sick of medical treatment between 3-6 months. It is almost similar to the study reported in Northeast Ethiopia in which nearly half (49.1%) of the cases had persistent lesions for less than six months before seeking medical care and in the University of Gondar, Ethiopia in which the median (IQR) duration of lesion before visiting the hospital was 12 (6-24) months [11, 14]. However, this finding is shorter than the report of the study conducted in Kenya in which the median duration of illness was 2years (range 1-4 years), and in ALERT Hospital, Addis Ababa in which most of the CL patients were visiting the hospital after 12 months following the appearance of the lesion [13,10].

This study revealed that there was a case build-up in July (6 cases), February (8 cases), March (8 cases), August (10 cases), and January (17 cases). About one-fourth (26.5%) of CL cases were reported in January, followed by August 10(15.63%). These are in line with the study conducted in Northeast Ethiopia, in which the cumulative monthly distribution of the disease was observed primarily in July (20.6%), June (14.2%), and January (11.7%); in Al Hassa, Saudi Arabia in which the number of cases showed a steep increase starting from November, reached a peak during January and February. The study conducted at the University of Gondar Hospital, Northwest Ethiopia, in which the highest prevalence rate (63.8%) was reported in September, followed by January (59.7%) and May (56.9%) [11,16,12].

The study showed that there were no cases reported in June and October. In contrast to this, the study conducted in Pakistan showed that the highest peak observed was in June and September and in Kenya, which revealed that there was an occasional case peak in June, and at the University of Gondar Hospital, Northwest Ethiopia, which showed that the least percentage (49.3%) was reported in June [23,13,12].

In this study, there was variation across the seasons in which more than half (51.56%) of CL cases were reported in winter (January and February), followed by summer (July and August) seasons (29.6%). In contrast to this, the study conducted in Larstan, South of Iran, showed that maxi-mum infection reports of 1,315 (26.06%) were in autumn in 2010, and the minimum infection reports of 160 (3.23%) were in winter of 2013 [20]. These could be explained by the fact that there might be differences in geographical characteristics and climatic conditions.

## Limitation of the study

As a limitation, the study was retrospective in design. Second, hospital records available for the review were either incomplete or inaccurate. As it was secondary data, essential variables such as environmental conditions, socioeconomic status, demographic, and human behaviours were not collected. These could lead to underestimating or overestimating the burden of cutaneous leishmaniasis in the study area. In addition to this types of lesions and sites of CL infection were not recorded. Finally, types of CL including diffused, localized or mucocutaneous were not assessed. The result should, therefore, be interpreted with consideration of the limitation.

## Conclusion and Recommendation

This study revealed that the trends of CL cases were fluctuating for the last more than three consecutive years (09 October 2018 to 31 January 2022) in the study area and continued as a public health problem in western Ethiopia. Those who live in rural areas, children and adolescents of school-going age were disproportionately affected. Hence, health care workers at different levels should give health education and/or health information to the rural and school communities in the study areas. This study revealed that there was a delayed seeking of medical treatment after being infected. Hence, health care professionals should give health education and promotion to improve the health-seeking behaviour of the communities. Besides, CL prevention and control interventions programs should be implemented by primary health care workers at grass root levels of the study areas in general and rural areas. Moreover, community-based research programs to determine the exact incidence and prevalence of CL cases and associated risk factors are needed.

## Data Availability

The finding of this study was generated from the data collected and analyzed based on stated methods and materials. The original data supporting this finding are available from the corresponding author on reasonable request.

## Acknowledgements

We are grateful to health care professionals and leaders of NSH; especially those working in a leishmaniasis treatment center for their valuable contribution during the study. Our special thanks also go to Sr. Tigist(Nurse in the leishmaniasis treatment center) and Dr. Getanu(Medical Director) of the hospital for facilitating the process of data collection.

## Lists of Abbreviations

ALERT: All African Leprosy Rehabilitation and Training Center
CL: Cutaneous leishmaniasis
NSH: Nekemte Specialized Hospital

## Declarations

### Ethics approval and consent to participate

Approval letter of permission was obtained from East Wollega Zonal Health Department, and the NSH medical director before the study was conducted to comply with the ethical guidelines. We used patient identifiable codes to maintain the confidentiality of each patient.

#### Consent for publication

‘Not applicable’

## Authors’ contributions

**ZK** participated in the design of the study, performed the data collection and statistical analysis, and served as the corresponding author of the manuscript. **YM** and **GM** supervised the study, ensured the quality of the data, assisted in the analysis and interpretation of the data. All authors read and approved the manuscript.

## Authors’ information

**ZK** is Public Health Specialist and Researcher at East Wollega Zonal Health Department, Nekemte, Western Ethiopia.

**YM** is WHE Public Health Emergency Surveillance Officer and Researcher, Nekemte, Ethiopia.

**GM** is Wollega University, School of Nursing and Midwifery, Department of Nursing, Nekemte, Ethiopia.

